# A study protocol for a retrospective cohort study and interrupted time series analysis to assess the effect of the COVID-19 pandemic on major trauma presentations and patient outcomes in English hospitals

**DOI:** 10.1101/2022.05.10.22274804

**Authors:** Carl Marincowitz, Omar Bouamra, Tim Coates, Dhushy Kumar, David Lockey, Lyndon Mason, Virginia Newcombe, Julian Thompson, David Yates, Fiona Lecky

**Affiliations:** Centre for Urgent and Emergency Care Research (CURE), Health Services Research School of Health and Related Research, University of Sheffield, Regent Court, 30 Regent Street, Sheffield, S1 4DA, UK; Trauma Audit Research Network, University of Manchester, Manchester, UK; Emergency Medicine Academic Group, Department of Cardiovascular Sciences, University of Leicester, University Road, Leicester LE1 7RH, UK; Department of Critical Care, Anaesthesia and Pre-hospital Emergency Medicine, University Hospital Coventry, Coventry, UK; London Air Ambulance, Royal London Hospital, Whitechapel Road, London, E1 1BB, UK and North Bristol NHS Trust, Southmead Way, Bristol, BS10 5NB, UK; Liverpool University Hospitals NHS Foundation Trust, University of Liverpool, Liverpool, UK; Division of Anaesthesia, University of Cambridge, Cambridge, UK; Department of Anaesthesia and Intensive Care Medicine, Southmead Hospital Intensive Care Unit, Southmead Hospital, North Bristol NHS Trust, Bristol, UK

## Abstract

A protocol for a retrospective cohort study and interrupted time series analysis to investigate the effect of successive COVID related “lockdown” restrictions on major trauma presentations and patient outcomes in English hospitals. The study specifically aims to assess: 1) The impact of successive “lockdowns” on the volume, demographics, injury mechanism, severity, treatment and outcomes of major trauma in England. 2) If the implementation of “lockdowns” affected major trauma related mortality.

A patient cohort will be derived from the Trauma and Audit Research Network (TARN) database, for all trauma receiving hospitals in England, between 1^st^ of January 2017 to 1^st^ of September 2021. This period encompasses two national “lockdown” periods (23^rd^ March 2020 to 29^th^ June 2020 and 2^nd^ Nov 2020 to 16^th^ May 2021) in England. A time series will be used to illustrate changes in the volume and mechanism of injury associated with successive “lockdowns”. Demographic characteristics and features of the clinical care pathways will be compared during the “lockdown” and equivalent pre-COVID periods. To specifically assess if there were any changes in risk adjusted mortality associated with the “lockdowns” interrupted time series analysis will be conducted.

## Background

To control transmission of COVID-19 during the pandemic the Government of the United Kingdom (UK) implemented successive “lockdown” measures in England.^1^ The devolved nations (Scotland, Wales and Northern Ireland) introduced similar measures but at different times. In England, the first “lockdown” was announced on the 23rd of March 2020 and, following a period of relaxation, a second “lockdown” was announced on the 30^th^ October due to the emergence of the Alpha variant. This led to a 16% reduction in road traffic, compared to an equivalent pre-pandemic period.^2^ This has been argued to present a unique opportunity to assess the impact of potential road traffic reducing public health measures on major trauma.^3^

However, there are also concerns that restrictions associated with “lockdowns” may have contributed to increased non-accidental injury, domestic violence and self-harm related to deteriorating mental health.^4-6^ Internationally, there is also evidence that despite “lockdown” measures, diversion of health care resources to treating patient with COVID-19, particularly intensive care capacity, may have led to worse outcomes for severely traumatically injured patients.^7^

Within the context of the UK there have been single centre or regional assessments of the impact of the first “lockdown” on the volume and characteristics of major trauma presentations.^8-11^ There has been no previous national evaluation of the impact of successive “lockdowns” on major trauma presentations and outcomes in the UK.

## Aims

To assess the impact of successive “lockdowns” on the volume, demographics, injury mechanism (particularly self-harm, interpersonal violence, other types of non-accidental injury and road traffic accidents), severity, treatment and outcomes of major trauma in England.

To assess if the implementation of “lockdowns” affected major trauma related mortality.

## Methods

### Study design

A retrospective cohort study and interrupted time series analysis using Trauma and Research Network (TARN) data for England.

### Data set

Routinely collected data for all TARN submitting hospitals in England for the period 1^st^ of January 2017 to 1^st^ of September 2021 will be used. This period encompasses two “lockdown” periods (23^rd^ March 2020 to 29^th^ June 2020 and 2^nd^ Nov 2020 to 16^th^ May 2021), a prolonged pre-COVID period to establish baseline trend and the period between the two “lockdowns”.

All trauma receiving hospitals (major trauma centres and trauma units) in England submit data on eligible trauma patients to the TARN database for the purposes of audit, governance and benchmarking.

The TARN database includes patients of any age who sustain injury resulting in: hospital admission > 72⍰h, critical care admission, transfer to a tertiary/specialist centre or death within 30⍰days. Isolated femoral neck or single pubic ramus fracture in patients > 65⍰years and simple isolated injuries are excluded. After study inclusion, a dataset of prospectively recorded variables covering demographics plus injury-related physiological, investigation, treatment and outcome parameters are collated using a standard web-based case record form by TARN hospital audit coordinators. Injury descriptions from imaging, operative and necropsy reports are submitted by TARN coordinators - all injuries are coded centrally using the Abbreviated Injury Scale, this enables calculation of the Injury Severity Score (ISS).

### Inclusion Criteria

All TARN eligible patients identified between the 1^st^ of January 2017 to 1^st^ of September 2021.

### Analysis

To illustrate changes in the volume and mechanism of injury associated with successive “lockdowns”, a quarterly time series for the period 1^st^ of January 2017 to 1^st^ of September 2021 will be conducted for the total volume of major trauma in England. The time series will be further stratified by management in a major trauma centre (MTC), road traffic accidents, intentional injuries (further stratified into types of interpersonal violence, paediatric non-accidental injury and self-harm) and other types of unintentional injury.

Demographic characteristics of TARN eligible patients including age, gender, physiology, injury severity and body region injury will be compared for the first “lockdown” (24th March to 3^rd^ July 2020 inclusive) and second lock down (1^st^ November 2020 to 16^th^ May 2021 inclusive) and will also be compared to equivalent pre-COVID-19 periods in 2018-2019. Similarly, to assess changes in management pathways for patients, the total and proportion of traumatically injured patients who were received by or transferred to a MTC, assessed by a consultant in the Emergency Department, received CT imaging, underwent an operation, were admitted ICU and or died in hospital will be compared in the pre-COVID and “lockdown” time periods.

To specifically assess if there were any changes in risk adjusted mortality associated with “lockdown” a weekly time series of risk adjusted mortality rate per 100, 000 will be plotted for the period 29^th^ October 2018 to 16^th^ May 2021. The weekly W statistics will be calculated for each consecutive weekly period using the conventional TARN method. The W can be interpreted as the number of excess survivors per 100 patients (observed – expected given case mix). Interrupted times series (ITS) analysis will be conducted to assess the impact of the “lockdowns” on the baseline trend of risk adjusted mortality. A segmented regression model predicting the weekly risk adjusted mortality will be estimated and a discontinuity in the gradient (trend) or intercept (level) of the fitted model will be tested for at the weekly time point of implementation of each “lockdown” (24^th^ March 2020 and 2^nd^ November 2020) and at the time of relaxation of the first “lockdown” (29^th^ June). Dates are chosen to incorporate the week each policy was implemented.

### Ethical, Regulatory Considerations and Dissemination

This analysis will be conducted on fully anonymised Trauma Audit and Research Network (TARN) data. The UK Health Research Authority Patient Information Advisory Group (PIAG) has given approval (Section 251) for analysis of anonymised Trauma Audit and Research Network (TARN) data. Details of the Section 251 approval are available here: https://www.tarn.ac.uk/Content.aspx?ca=2&c=3857.

We intend to present the results of this study at conferences and to publish the results in a peer-reviewed scientific journal.

### Study Team

The research team includes a range of clinical and statistical methods experts on the evaluation of trauma care in the UK. FL and TC are professors of Emergency Medicine and DSK is a consultant in critical care who have collaborated on a range of TARN and other trauma related clinical trials and other research projects. They have permanent positions in TARN (FL is Research Director, DSK is director of clinical audit and TC is chairman of the TARN executive board). OB is the TARN permanent medical statistician. DY is a professor emeritus of Emergency medicine and member of the TARN research committee. CM is an NIHR Clinical Lecturer in Emergency Medicine and member of the TARN research committee co-ordinating this project.

External investigators providing additional expertise include: DJL intensive care consultant and Hon. Professor of Trauma & Pre-hospital Emergency Medicine, JT intensive care consultant and Major Trauma Network Research Lead. LM associate professor and consultant in Trauma and Orthopaedics and VN is an Honorary Consultant in Neurosciences and Trauma Critical Care and Emergency Medicine and Royal College of Emergency Medicine Associate Professor.

### Risks and anticipated benefits for trial participants and society

The study will use existing fully anonymised data derived from clinical routinely collected for the purposes of audit of trauma care and not alter patient management. The risks to patients involved in the study are therefore very low and principally relate to data protection and confidentiality.

Future trauma patients and society in general will benefit from an evaluation of whether changes in behaviour as result of “lockdown” measures affected presentations and outcomes related to major trauma. The results may also inform future NHS decision making about the balance between continuing routine care and responding to public health emergencies. Such information could also inform public health interventions aimed at reducing trauma related to road traffic accidents and non-intentional injury.

## Data Availability

Data may be obtained from a third party and are not publicly available. The de-identified patient data used for this study are the property of the Trauma Audit and Research Network (TARN), based at the University of Manchester. These data may be requested directly from TARN.

